# Stress-mediating Inflammatory Cytokine Profiling Reveals Unique Patterns in Malaria and Typhoid Fever Patients

**DOI:** 10.1101/2024.06.23.24309363

**Authors:** MacDonald Bin Eric, Palmer Masumbe Netongo, Severin Donald Kamdem, Nzuno Christine, Tchoutang Ange Maxime, Tchoupe Kamoua Eric Berenger, Bongkiyung Donald Buri, Ngum Leslie NGum, Jean Paul Chedjou, Akindeh Mbu Nji, Wilfred Fon Mbacham

## Abstract

**Background:** Malaria and typhoid fever are known to cause severe morbidity and mortality if miss-treated or left untreated. The oxidant/antioxidant and immune responses to these diseases involve a complex interplay of stress-mediating inflammatory cytokines which orchestrate a coherent response that leads to pathogen clearance and recovery. This study investigated the stress-mediated cytokine profiles in patients attending the Obala District Hospital, Yaoundé, Cameroon and diagnosed with malaria and/or typhoid fever.

**Methods:** In this cross-sectional observational study, we measured the levels of cortisol and stress-mediating inflammatory cytokines in peripheral blood samples collected from 55 malaria and/or typhoid fever patients, along with a healthy control group (n = 15) using commercial ELISA kits. The psychological stress of the voluntary participants over the past 30 days was assessed using a 10-item Perceived Stress Scale (PSS) questionnaire.

**Results:** Compared to the control group, patients with co-infection showed significant differences in anemia, thrombocytopenia, and monocytosis. Psychological stress levels were insignificantly higher in patients with typhoid fever (18.20±5.5) compared to control (15.0±2.43), malaria (15.80±4.39), and comorbidity (15.40±5.26) groups. Cortisol levels were significantly (p<0.001) higher in all patient groups compared to the control group, with the typho-malaria group showing a 2.5-fold increase. Cytokine levels were also elevated in patients with malaria and typhoid comorbidity, particularly IL-1β, IL-2, TNF-α, and IFN-γ. While IL-6 concentrations were significantly higher in patients with malaria and typho-malaria comorbidity, IL-10 levels were reduced in the typho-malaria group but still higher than in the control group. The TNF-α/IL-10 ratio was significantly higher in the co-infected group, indicating a more intense inflammatory response. Interestingly, there was a positive correlation between perceived stress scores and IL-2 (r = 0.365, p=0.002), IFN-γ (r = 0.248, p=0.03) and IL-6 (r = 0.412, p=0.0001) in the typho-malaria group. Beside IL-6, no significant correlation was found between stress index and the other anti-inflammatory cytokines IL-4 (r = 0.204, p=0.09) and IL-10 (r = 0.153, p=0.20) in coinfected individuals.

**Conclusion:** These results suggest that stress response may play a crucial role in shaping the inflammatory landscape during malaria and typhoid fever. Exposed to severe stressors may disrupt immune response and contribute to negative health outcomes. Understanding the immunopathogenesis of these diseases could potentially pave the way for the development of novel therapeutic strategies targeting the stress-cytokine axis.

## Introduction

Malaria is a serious disease that affects millions of people worldwide and is caused by parasites transmitted through the bite of *Plasmodium*-infected mosquitoes. According to recent data from the World Health Organization, there were an estimated 249 million malaria cases in 2022, resulting in approximately 608 000 deaths (1). It remains a leading cause of morbidity and mortality in many parts of the world. The *Salmonella enterica serotypes Typhi, Paratyphi A, B*, and *C* cause potentially severe and occasionally life-threatening bacteremic illnesses collectively referred to as enteric fever. Typhoid fever outbreaks have been reported in many developing countries, particularly in areas with large populations and inadequate health infrastructures (2). In addition, the emergence of antibiotic-resistant strains of *Salmonella* species has made controlling the disease more challenging. WHO in 2021 estimates about 11–21 million cases of typhoid fever and 5 million cases of paratyphoid fever worldwide each year, causing an estimated 135 000 – 230 000 deaths.

Malaria and typhoid fever are two of the most common infectious diseases that impact a significant portion of the global population including Cameroon. The burden of these diseases includes not only the physical suffering experienced by those infected with the disease but also the economic and social impacts on the communities affected, particularly in developing countries where healthcare infrastructures are often limited and are characterized by poor sanitation and limited access to clean water or food. For individual patients, both malaria and typhoid fever typically cause a wide range of overlapping symptoms, including fever, headache, muscle aches, fatigue, and abdominal pain (3). In rare cases, patients may develop life-threatening complications. This homology in clinical feature of the two diseases pushes individuals who practice symptom-base treatment without prior diagnosis to treat one disease for the other (4).

Cytokines, as key components of the immune system, play a crucial role in the body’s response to malaria and typhoid fever. However, the immunopathogenesis of these diseases is still not well elucidated. During *Plasmodium* infections, pro-inflammatory cytokines like tumor necrosis factor (TNF), interleukin-1 (IL-1), and interleukin-6 (IL-6) are released from immune cells in response to the parasite antigens (5,6). Elevated levels of these cytokines have been associated with the severity of malaria symptoms and complications like cerebral malaria (5). TNF in particular has been shown to mediate pathological effects during severe malaria by increasing vascular permeability, activating endothelial cells, and promoting leukocyte adhesion (7). For typhoid fever, pro-inflammatory cytokines including IFN-γ, IL-6, IL-10, TNF-α and TNF are also produced in large amounts (8,9). High concentrations of these cytokines have been linked to typhoid fever complications (10). Thus, an excessive inflammatory response mediated by cytokines like TNF, IL-1, and IL-6 likely contributes to the tissue damage and organ dysfunction observed in severe malaria and typhoid fever.

A recent study demonstrated a fundamental relationship between the immune and endocrine systems to modulate an adequate response to physiological and psychological stressors (11). Although the exact mechanisms governing the interplay between cortisol and cytokines are still not fully elucidated, the reciprocal interaction between cortisol and inflammatory cytokines is crucial for maintaining the delicate balance between the hypothalamic–pituitary–adrenal (HPA) axis and the immune system, ensuring optimal functioning of both systems during infection. This study was designed to investigated the effect of malaria and typhoid comorbidity on stress-mediated cytokine concentration in patients diagnosed with typhoid fever and/or malaria.

## Research methods

### Study design

We conducted a cross-sectional observational study involving patients diagnosed with malaria and/or typhoid fever seeking medical attention at the Obala District (rural) hospital in Yaoundé, Cameroon from September, 2022 to June, 2023.

### Sample size

In order to balance between study feasibility and statistical power, minimise risk of dropouts, a convenient sample size of 70 participants (55 malaria and/or typhoid fever confirmed cases and 15 healthy controls) were recruited.

### Patient Selection and clustering

We included voluntary participants of age 10 years and above, who provided written informed consent and assent. The patients were selected based on their infectious status of either malaria or typhoid fever (**Figure 1**). A stratified random sampling technique was used where the population was divided into four groups (strata): Malaria (Mal^+^), Typhoid (Typ^+^), (TypMal^+^) and Healthy control (TypMal^-^). Individuals with similar characteristics suffering from different diseases were paired to help control for confounding variables. We excluded pregnant women, patients with severe complications of malaria or typhoid fever, HIV-positive patients, other comorbidities such as diabetes, hepatitis. These conditions can influence the stress/immune responses in patients with typhoid fever and/or malaria by altering the disease severity.

**Figure 1:**
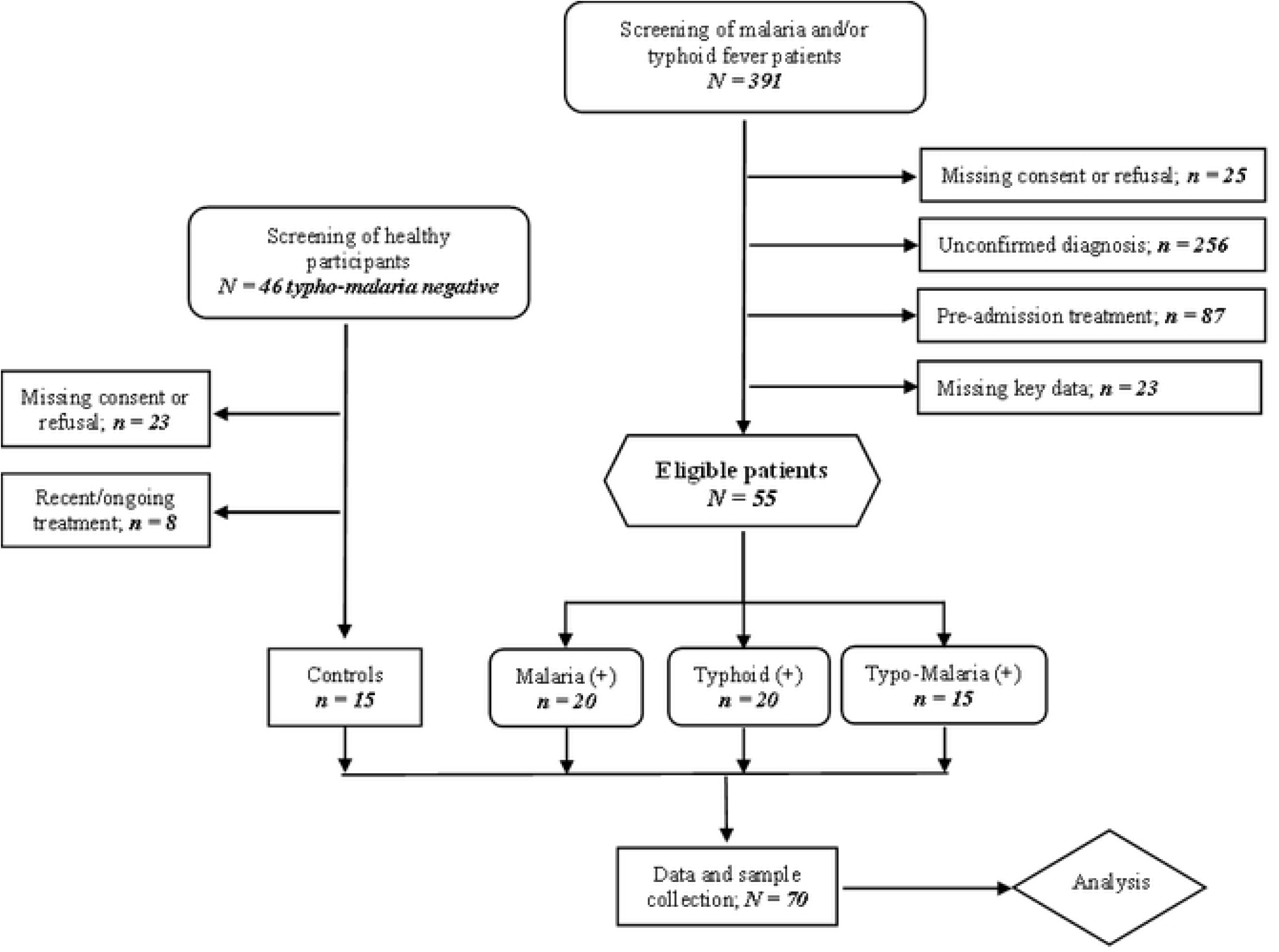
The study flowchart indicating the strategy of selection and clustering of participants.

### Data collection

Participants’ data were collected by trained clinical personnel using a standardized case report form (CRF). Data collected included demographic information, patient history, symptoms & signs (type and severity), co-morbidities, initial diagnosis, drugs and test prescribed, laboratory results and care pathway (hospitalization or outpatient care).

### Stress event measurements

We collected information about stressful life events which had occurred in the previous month using a 10-item Perceived Stress Scale (PSS) (12) over the past 30 days on a 5-point scale (0 = never, 1 = almost never, 2 = once in a while, 3 = often, 4 = very often). A perceived stress score ranging from 0-13 was considered “low stress”, 14-26 as “moderate stress” and 27-40 as high perceived stress.

### Sample collection and laboratory analysis

Patients with signs and symptoms presumptive of malaria and/or typhoid fever were clinically screened and suspected participants were prescribed both malaria and typhoid fever tests. After the patient had been clinically diagnosed for malaria and/or typhoid fever, about 15 ml of blood sample were collected. Blood samples were then distributed in EDTA, dry and blood culture tubes. All laboratory analyses were performed by competent laboratory personnel.

### Diagnosis and confirmation of *Plasmodium* infection

Malaria diagnosis was done using standard diagnostic methods as described elsewhere (4). Malaria Rapid Diagnostic Tests were used to detect the presence of *Plasmodium* antigens in the patient’s blood in accordance with the manufacturer’s instructions. Thick and thin blood films were also prepared and stained with Giemsa for a confirmatory diagnosis of malaria and parasite count using microscopy. Two experienced laboratory personnel independently examined the slides for the presence of *Plasmodium* parasites.

### Diagnosis and confirmation of typhoid fever

#### Rapid diagnosis of typhoid fever

Package inserted protocol instructions were followed. The laboratory screening of typhoid fever was done using the OnSite Typhoid IgG/IgM Combo rapid test (CTK Biotech Inc, USA), a lateral flow chromatographic immunoassay that detects and distinguishes IgG and IgM antibodies to *Salmonella typhi* and *S. paratyphi* in human blood. Results were interpreted as described elsewhere (13).

#### Widal agglutination test

Widal test was performed on acute serum using Sanofi qualitative agglutination test kits (Bio-Rad) containing coated somatic (O) and flagella (H) antigens of *S. typhi* and *S. Paratyphi A, B* and *C*. Analysis was initially carried out as described previously (4). All positive results obtained through a slide test were confirmed and quantify by the tube agglutination method. A positive Widal test was considered as one that gave a reaction titre greater or equal to 1/200 for *Salmonella* somatic and flagella antibodies, after the prescribed incubation time (14). However, the last test showing signs of agglutination was taken as the titre for that patient (15).

#### Culture and identification of *Salmonella spp*

About 4 ml of venous blood sample collected from each participant was immediately inoculated into bottle containing 46 ml triptic soya broth medium (Himedia, India) and incubated for 7 days. The cultured bottle which showed growth were further sub cultured on MacConky agar (Deben diagnostic Ltd) and blood agar media (Biomark, India laboratories) after 48 h. Negative broth culture were incubated for seven days and sub cultured before reported negative. Specific antisera were used to determine *Salmonella spp*.

### Estimation of inflammatory cytokines levels

Blood samples were transferred to the Molecular Diagnostic Research (MDR) Laboratory (Yaoundé, Cameroon) and Laboratory for Public Health Research Biotechnologies (LAPHER BIOTECH) at the Biotechnology Centre (University of Yaoundé I, Cameroon) within 2 h, in cooled bags, then centrifuged for 10 min at 3500 rpm at 4 °C to allow plasma and serum collection. Seven inflammatory cytokines were assayed in duplicate using ELISA: interleukin (IL)-1β (Cat #: EKHU-0083, Melsin, China), IL-2 (Cat #: EKHU-0144, Melsin, China), IL-4 (Cat #: EKHU-0014, Melsin, China), IL-6 (Cat #: EKHU-0140, Melsin, China), IL-10 (Cat #: EKHU-0155, Melsin, China), tumor necrotic factor (TNF)-α (Cat #: EKHU-0110, Melsin, China), and interferon (IFN) -γ (Cat #: EKHU-1695, Melsin, China).

### Assessment of stress response

The level of cortisol was estimated using commercially available enzyme-linked immunosorbent assay (ELISA) kit (Cat #: ARG8139, Arigo BioLaboratories, Taiwan). All blood parameters were tested following the manufacturer’s instructions.

### Ethical considerations

This study was conducted in compliance with the national and international ethical standards for research involving human participants. The study protocol was reviewed and approved by ethical review committee and regulatory authorities: the Centre Regional Ethics Committee for Human Health Research (Ref. #: 0226/CRERSHC/2022) and the Centre Regional Delegation of Public Health under the ministry of Public Health in Cameroon (Ref. #: 1392/AAR/MINSANTE/SG/DRSPC). Written informed consent/assent was obtained from all participants before enrollment, confidentiality and privacy was maintained, and any potential risks or adverse effects were greatly minimized.

### Data analysis

Statistical analysis was performed using SPSS (V.26) and GraphPad (Prism Software 9.0.0). Cytokine levels were compared among the groups. Statistical analyses were performed to determine the significance of any observed differences. Comparisons between the groups were performed with ANOVA as appropriate for continuous variables, and with chi-square test or Fisher exact test as appropriate for categorical variables. The changes in cytokine levels were then correlated with other clinical parameters. The differences were considered statistically significant if p<0.05.

## Results

### Assessment of general demographic characteristics of the study population

The general demographic characteristics of patients (n=55) and healthy controls (n=15) are presented in Table 1. Among eligible febrile patients, the ratio of females is to males was 1.04, with a mean (±SD) age of 25.06 ( ± 11.56) years; whereas among controls the ratio of females is to males was 1.5, with a mean (±SD) age of 30.60 ( ± 12.19) years. Aside from fever, the most frequently occurring symptoms were headache (54.55%), dizziness (49.09%) in study population. The mean time from onset of symptoms to admission was highest in typhoid fever mono-infections, 4.95 (±2.54) days. On admission, the mean (SD) perceived stress of the coinfected group was about one and a half times greater than the control group and almost similar to the typhoid fever group with no significant difference between groups.

**Table 1:** Baseline Characteristics of the study population.

| *Characteristics* |  | **Healthy control** | **Malaria mono** | **Typhoid mono** | **Typho-malaria** | *p-value* |
| --- | --- | --- | --- | --- | --- | --- |
| *(N = 70)* |  | *(n = 15)* | *(n = 20)* | *(n = 20)* | *(N = 15)* |  |
| *Mean age (±SD)/years* |  | 30.60 (± 12.19) | 22.35 (± 14.54) | 27.50 (± 9.50) | 25.33 (±10.65) | 0.228 |
| *Gender* | Male, **n (%)** | 6 (40.00) | 12 (60.00) | 7 (35.00) | 8 (53.33) | 0.211 |
|  | Female, **n (%)** | 9 (60.00) | 8 (40.00) | 13 (65.00) | 7 (46.67) |  |
| *Core temperature (SD)/°C* |  | 36.7 (±0.38) | **38.44 (±0.67)\*** | **38.55 (±1.09)\*** | **39.07 (±1.18)\*** | **<0.001** |
| *Clinical presentation on admission* |  |  |  |  |  |  |
| *Headache, n (%)* |  | / | 11 (55.00) | 8 (40.00) | 11 (73.33) | 0.224 |
| *Anorexia, n (%)* |  | / | 9 (45.00) | 5 (25.00) | 7 (46.67) | 0.157 |
| *Sweating, n (%)* |  | / | 2 (10.00) | 3 (15.00) | 4 (26.67) | 0.212 |
| *Dizziness, n (%)* |  | / | 8 (40.00) | 9 (45.00) | 10 (66.67) | 0.084 |
| *Body pain, n (%)* |  | / | 7 (35.00) | 4 (20.00) | **10 (66.67)\*** | **0.024** |
| *Vomiting, n (%)* |  | / | 4 (20.00) | 2 (10.00) | 3 (20.00) | 0.131 |
| *Diarrhea, n (%)* |  | / | 4 (20.00) | 3 (15.00) | 9 (60.00)* | **0.029** |
| *Abdominal pain, n (%)* |  | / | 2 (10.00) | 6 (30.00) | 8 (53.33) | 0.214 |
| *Anaemia, n (%)* |  | / | **6 (30.00)\*** | 2 (3.64) | **6 (10.91)\*** | **<0.001** |
| *Mean onset of symptoms (SD)/days* |  | - | 2.10 (±1.55) | 4.95 (±2.54) | 4.00 (±1.73) | 0.147 |
| *Average weight (SD)/kg* |  | 64.64 (±10.59) | 50.97 (±19.19) | **64.34 (±10.57)\*** | 57.46 (±13.33) | **0.042** |
| *Mean systolic BP (SD) /mmHg* |  | 115.53 (±3.85) | **108.80 (±6.46)\*** | **109.60 (±8.04)\*** | 114.07 (±6.97) | **<0.01** |
| *Mean diastolic BP/mmHg* |  | 65.73 (±2.96) | **59.85 (±5.95)\*** | 64.35 (±5.81) | 62.80 (±8.11) | **0.027** |
| *Median parasite density (range)/µl* |  | / | 1850.00<br>(105 - 103025) | / | **12850**<br>**(1035 – 122045)\*** | **<0.001** |

*Data represented as count (percentage, %), mean (standard deviation) and range (min. - max.) where the asterisk (\*) corresponds* *to statistical significance at p<0.05.*

### Laboratory findings

#### Hematological parameters

The results of hematological parameters are shown in **Table 2**. Malaria patients in this study showed a mild state lymphocytopenia with less than 1000 lymphocytes per microliter of blood. Anemia, thrombocytopenia and monocytosis were equally recorded in the co-infected groups with significant difference from the control group.

**Table 2:** Hematological parameters associated with malaria, typhoid and typho-malaria comorbidity.

| *Blood parameter* | *Control*<br>*n=15* | *Malaria mono*<br>*n=20* | *Typhoid mono*<br>*n=20* | *Typho-malaria*<br>*co-infection,*<br>*n=15* | *p-value* |
| --- | --- | --- | --- | --- | --- |
| *Leukocyte (SD) x10<sup>9</sup>/L* | 4.51 (±0.51) | **5.08 (±0.78)\*** | 4.17 (±1.03) | 4.46 (±1.15) | **0.021** |
| *Hematocrit, (SD) %* | 43.78 (±2.70) | 40.8 (±2.75) | 41.9 (±2.36) | **38.47 (±3.54)\*** | **<0.001** |
| *Neutrophils (SD) x10<sup>9</sup>/L* | 1.97 (±0.24) | 2.13 (±0.37) | 1.95 (±0.29) | 1.84 (±0.37) | 0.079 |
| *Lymphocytes (SD) x10<sup>9</sup>/L* | 1.52 (±0.50) | **0.95 (±0.27)\*** | 1.19 (±0.35) | **0.98 (±0.21)\*** | **<0.001** |
| *Monocytes (SD) x10<sup>9</sup>/L* | 0.63 (±0.33) | 0.82 (0.48) | 0.99 (±0.48) | **1.15 (±0.37)\*** | **0.009** |
| *Eosinophils (SD) x10<sup>9</sup>/L* | 0.20 (±0.12) | **0.077 (±0.04)\*** | 0.11 (±0.06) | **0.09 (0.06)\*** | **<0.001** |
| *Platelets (SD) x10<sup>9</sup>/L* | 158.33 (±28.74) | 135.15 (±19.48) | 132.6 (±25.01) | **113.4 (±14.2)\*** | **<0.001** |
| *Hemoglobin (SD), g/dL* | 14.22 (±1.68) | 12.11 (±1.61) | 12.46 (±1.50) | **10.81 (±1.34)\*** | **<0.001** |

*Data represented as mean (standard deviation) where the asterisk (\*) corresponds to statistical significance.*

#### Stress response in the study groups

The stress response was evaluated by estimating the cortisol within the study population and the result are shown in **Figure 2A** below. The overall mean baseline cortisol levels were significantly higher in all patient groups especially in typho-malaria group, 43.27 (±8.12) compared to the malaria group, 38.39 (±11.80), typhoid fever group, 33.70 (±11.83) control, 17.01 (±11.30).

**Figure 2:**
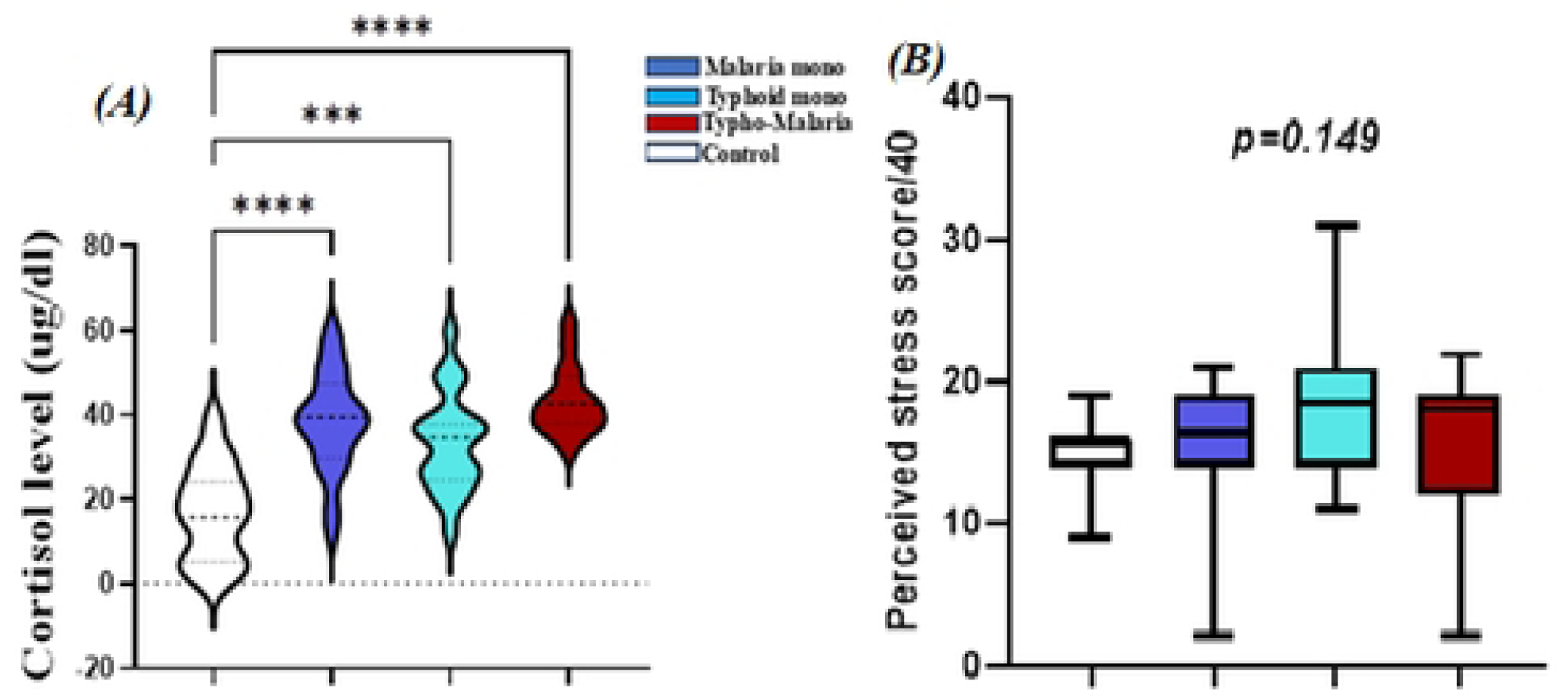
Comparison of cortisol levels and perceived stress score in the study groups *Data represented as concentration of cortisol in µg/dl and psychological stress score on a scale of 40 measured on admission prior to treatment and the asterisks denote a statistical significance at (*): p<0.05 [0.01 – 0.01]; (**\*\*\***): p<0.001 [0.0001 – 0.001] and (**\*\*\*\***): p<0.0001*.

Psychological stress results (**Figure 2B**) showed that patients with typhoid fever reported an insignificantly (p > 0.05) higher perceived stress levels (18.20±5.5) compared to the control group (15.0±2.43), patients with malaria (15.80±4.39), and comorbidity (15.40±5.26). Thematic analysis of the qualitative data revealed that patients with typhoid fever experienced more anxiety and fear related to the possibility of complications and treatment failure.

#### Immunological features of the study groups

On admission, the concentrations of IL-1β (*p<0.01*), IL-2 (*p<0.001*), TNF-α (*p<0.001*), and IFN-γ (*p<0.001*) were significantly upregulated in patients with malaria and typhoid comorbidity, comparing to healthy controls (**Figure 3**). Aside from the significant difference in IL-6 level recorded in the malaria mono and typho-Malaria groups (p<0.01), there were no statistically significant variation in IL-6 levels within the other groups (**Figure 3c**).

**Figure 3:**
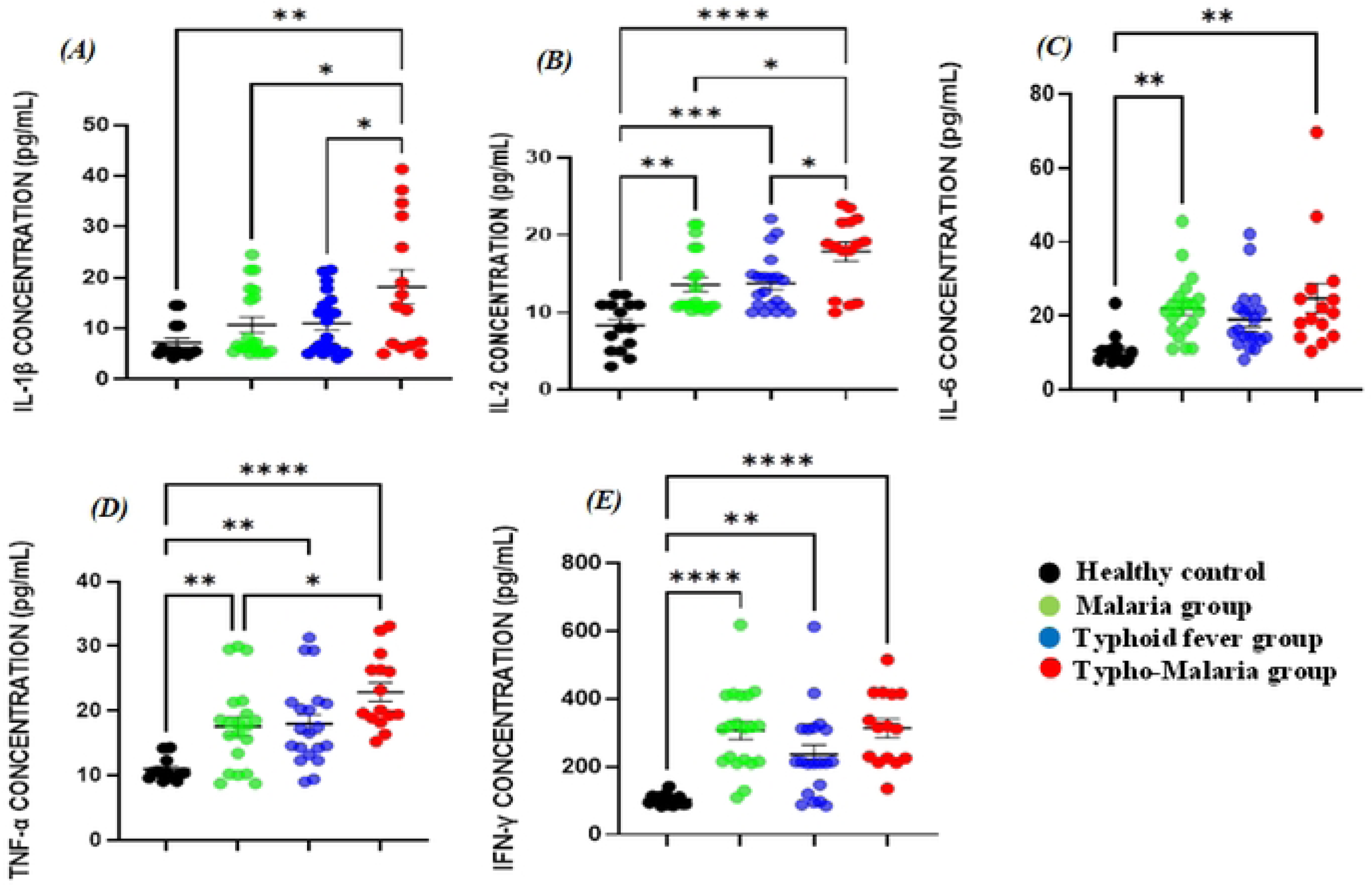
Comparison of pro-inflammatory cytokine levels in the study groups: **(A)** IL-1β; **(B)** IL-2; **(C)** IL-6; **(D)** TNF-α and **(E)** IFN-γ among the different groups: Malaria^+^ (n = 20), Typhoid^+^ (20) Typo-Malaria^+^ (n = 15) and Control (n = 15).| *Data represented as concentration in pg/ml measured on admission prior to treatment and the asterisks denote a statistical significance at (**\***): p<0.05 [0.01 – 0.01]; (**\*\***): p<0.01 [0.001 – 0.01]; (**\*\*\***): p<0.001 [0.0001 – 0.001]; and (**\*\*\*\***): p<0.0001*.

Anti-inflammatory cytokine levels on admission varied significantly across the groups (**Figure 4**). Generally, IL-4 concentrations were significantly higher in the Typhoid mono and Typho-Malaria groups compared the control (**Figure 4a**). On the other hand, IL-10 levels were slightly reduced in the Typho-Malaria group (255.3±177.0) (**Figure 4b**) compared to the mono-malaria (293.1±106.8) and typhoid fever (2262.6±131.9) groups, but significantly higher when compared with control (130.9±51.56). In spite the insignificant variation among groups, the malaria group showed a very strong significant rise in IL-10 (p<0.001) compared to the heathy control.

**Figure 4:**
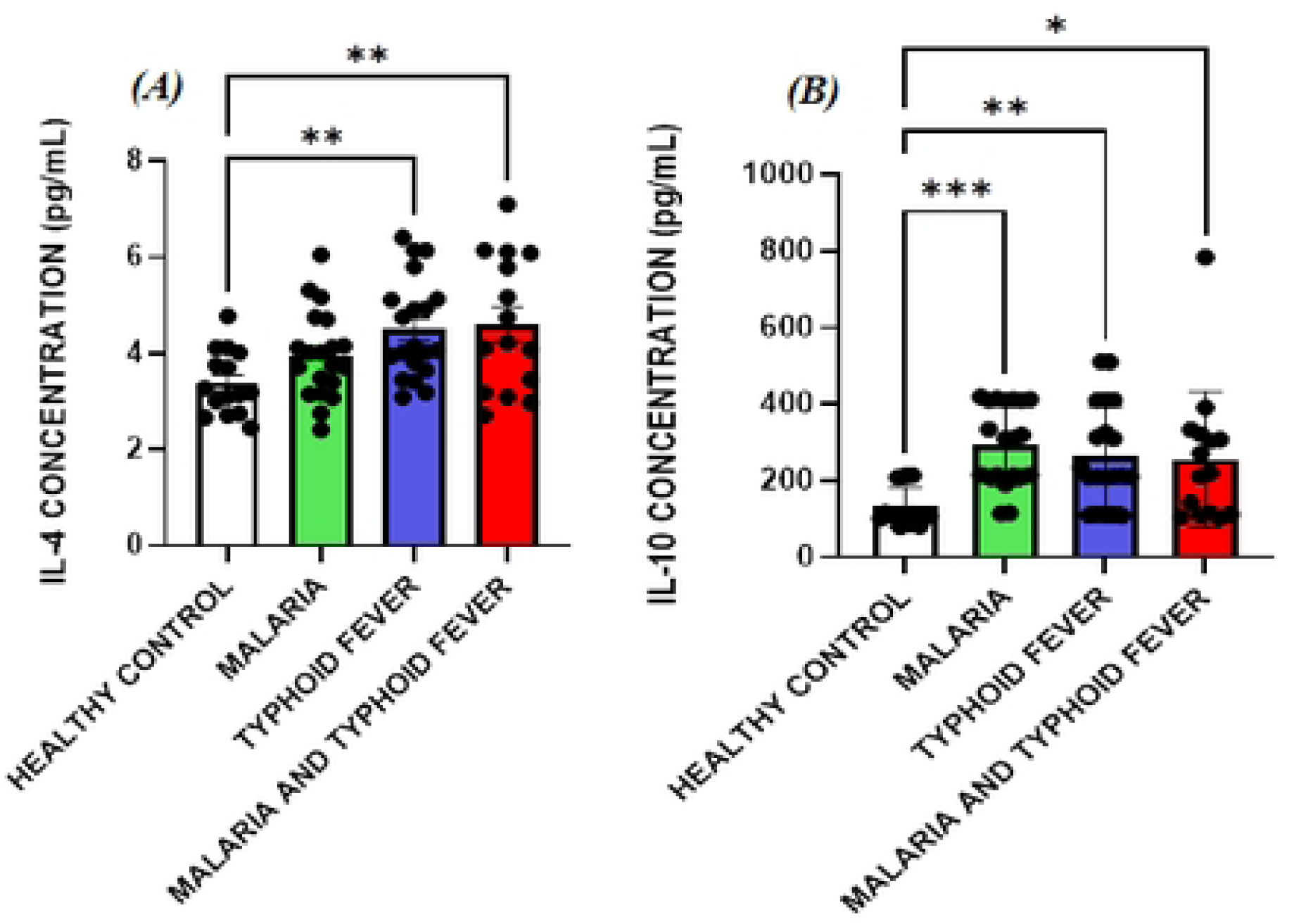
Comparison of anti-inflammatory cytokine levels in the study groups: ***(A)*** IL-4 and ***(B)*** IL-10: Malaria^+^ (n = 20), Typhoid^+^ (20) TypoMalaria^+^ (n = 15) and Control (n = 15). *Data represented as concentration in pg/ml measured on admission prior to treatment and the asterisks denote a statistical significance at (*): p<0.05 [0.01 – 0.01]; (**): p<0.01 [0.001 – 0.01]; (**\*\*\***): p<0.001 [0.0001 – 0.001]*.

#### Pro- and anti-inflammatory ratio compared

Disbalances in pro- and anti-inflammatory cytokines were observed in the study population. Therefore, we chose pro-inflammatory cytokines (IL-2, TNF-α and IFN-γ) which act as key actors in the regulation of infections with statistical significance (*p<0.001*) and divided the expression by IL-10 which was the key anti-inflammatory cytokine identified in the study group (*p<0.05*) to calculate a pro- and anti-inflammatory cytokine expression ratio. Results of these ratios where equally compared between groups as shown in **Figure 5**.

**Figure 5:**
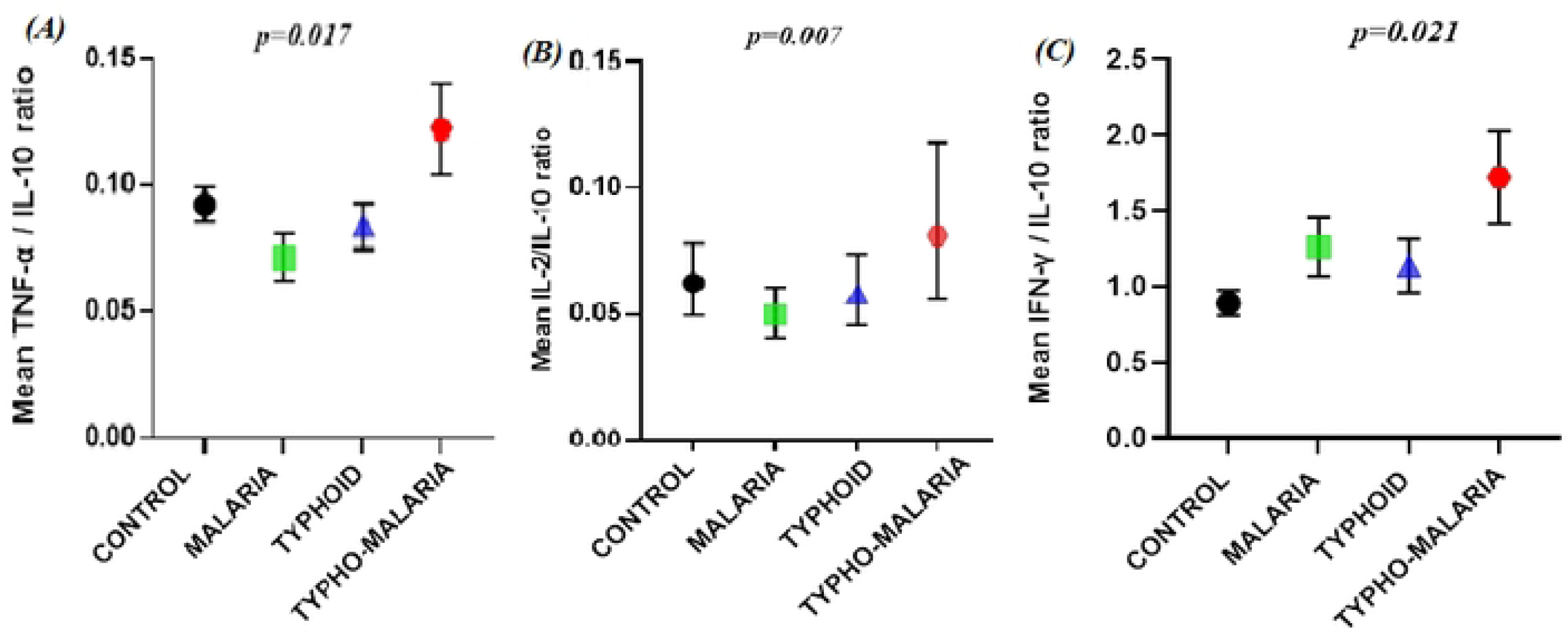
Comparison of the pro/anti-inflammatory cytokine ratios in the study groups: ***(A)*** TNF-α/IL-10; ***(B)*** IL-2/IL-10; ***(C)*** IFN-γ/IL-10. Data are presented as mean plots and p-values are derived from Mann-Whitney U test.

## Discussion

The pathogeneses of malaria and typhoid fever are quite complex and incompletely elucidated. In the present study, we investigated the interplay between stress response and inflammatory cytokine profiles in patients with malaria, typhoid fever, and co-infection with both diseases. Our findings revealed distinct clinical and blood parameters among the patient groups compared to healthy participants. There was no significant difference in age and gender between the study groups. the absence of significant differences in age and gender between the study groups minimizes bias and enable us to attribute observed differences to the pathogens rather than extraneous variables, which is essential for understanding the interplay between stress response and inflammatory cytokine profiles in patients with these diseases.

Results of this present investigation revealed, highly significant differences in core temperature (p<0.001), systolic (p=0.007) and diastolic (p=0.027) blood pressure, and most of the hematological parameters (**Table 2**). These findings suggest that the pathophysiological mechanisms of *Plasmodium* and *Salmonella spp in* co-existence may trigger distinct thermoregulatory patterns and different levels of cardiovascular instability which could be related to severity. Malaria patients in this study showed a mild state lymphocytopenia with less than 1000 lymphocytes per microliter of blood. Anemia, thrombocytopenia and monocytosis were equally recorded in the co-infected groups with significant difference from the control group. Leukopenia, anemia, and thrombocytopenia have been reported to be significantly associated with malaria and estimated to be specific for the diagnosis of malaria (16–19). The cause of thrombocytopenia in malaria and typhoid fever is poorly understood but, platelet indices could be useful predictors of disease severity (20,21). Correlation analysis revealed that duration of pre-admission symptoms in typho-malaria showed strong positive correlation with diastolic blood pressure (*r=0.421, p=0.001*) and Perceived Stress index (*r=0.369, p = 0.006*). This implies that prolonged exposure to the pathogens may lead to increased psychological and cardiovascular stress, which could trigger or worsen hypertension and exacerbate the physical symptoms. This highlights the importance of considering the psychological toll and duration of symptoms when evaluating patients with malaria and/or typhoid fever.

Additionally, in the typho-malaria group, perceived (psychological) stress index gave strong positive correlation with parasite density (*r=0.613, p<0.001*) and eosinophil count (r=0.303, p=0.01) while core temperature and eosinophil count were negatively correlated (r=-0.343, p=0.004). This implies that stress may play a role in modulating the immune response to infection during co-infections, potentially allowing for greater parasite growth. Eosinophils are a type of white blood cell that play a role in fighting parasitic infections. Thus, the decreased eosinophil counts recorded in patients with malaria and typho-malaria is likely to be associated with an impaired immune response, reduced inflammatory response, and increased risk of severe disease during plasmodium infection with less effect during typhoid fever. This finding highlights the importance of considering the psychological factors that may influence stress and immune response in patients with malaria.

An approximately three-fold increase in parasitemia in typho-malaria group compared to those patients with malaria only (geometric mean parasitaemia of 34711.2 versus 12744.1 per µl of blood, p value of <0.001) was observed. A similar significantly high parasitemia was reported previously by Netongo *et al*., in 2022 (4). This implies that co-infection may enhance the replication of the malaria parasites, impaired immune response or suppression of immune cells by the salmonella bacteria or other factors, and a potential for increased transmission. This highlights the importance of simultaneous diagnosis of both malaria and typhoid fever. Correlation analysis revealed a significant negative correlation between parasite density and hemoglobin count in both the malaria mono (r =-0.356, p=0.005) and coinfected groups (r=-0.457, p<0.001). This is consistent with the known effects of *Plasmodium* parasites on red blood cell production and destruction. It could be suggested here that the *Salmonella* organism contributes to parasite-mediated hemolysis and disruption of erythropoiesis, consistent with the known immunosuppressive effects of malaria on the bone marrow (22). Thus, by monitoring parasite density and hemoglobin levels, clinicians may be able to identify patients at high risk of developing anemia and provide targeted interventions earlier. A similar result was recently reported in malaria patients by Antwi-Bafour *et al*., 2023 (23).

For a better understanding of the physiological stress response to infection in patients with malaria and typho-malaria, we measured the concentration of cortisol in all study groups. The baseline cortisol levels were significantly higher in all patient groups when compared to the control (p<0.001). The overall mean cortisol level in the typho-malaria group was over 2.5 times greater than that of the control (43.27 versus 17.01). This is contrary to the report of Ibrahim EA et al., in 2011 (24) who recorded no significant difference in mean cortisol levels in patients with malaria in comparison with the control group. It could thus be suggested that the stress response is activated in patients with malaria and typhoid fever and in an attempt to cope with this threat, a physiological response is triggered. Also, elevated cortisol levels may indicate dysregulation of the hypothalamus/pituitary/adrenal axis (HPA) axis, which is responsible for regulating the body’s response to stress. This can favour the development of complications and/or contribute to treatment failures. Thus, clinicians may need to consider the possibility of hypercortisolism when managing patients with these conditions as medications that target the HPA axis could be used to regulate cortisol levels and alleviate symptoms. Elevated cortisol levels have been attributed to increased risk of complications in recent studies, such as hyperglycemia (25), hypertension (26), and impaired immune function (27).

Correlation analysis revealed that rise in serum cortisol level had a positive correlation with core temperature in all patient group with the strongest correlation in the typho-malaria group (r=0.492, p<0.0001) and thus can be useful to predict the severity in malaria and typhoid fever comorbidity. A current investigation confirmed that temperature interfered with the cortisol secretion; suggesting stimulation of this hormone in malaria patients (28). On the other hand, cortisol levels showed a strong negatively correlation with hemoglobin (r=-0.352, p=0.003) and platelet counts (r=-0.373, p=0.02) in the typho-malaria group, with an insignificant positive correlation with leukocyte (r=0.144, p=0.23). This finding implies that cortisol levels may be contributing to the hemolytic anemia and thrombocytopenia commonly seen in patients with malaria and typhoid fever. In the malaria mono group, cortisol levels were equally negatively correlated with lymphocyte counts (r=-0.336, p=0.004). This could be due to the immunosuppressive effects of cortisol, which can inhibit the activity of lymphocytes and other immune cells. Contrarily, in the typhoid mono group, there was a weak positive correlation between cortisol and leukocyte counts (r=0.312, p=0.04) which could be due to the stimulatory effects of cortisol on the bone marrow, which can lead to increased production of leukocytes. However, reports highlighting the role of cortisol in typhoid fever pathogenesis are scarce.

It has been documented that the blood-stage cycle of the malaria parasite is characterized by an upregulation of inflammatory cytokines like IL-6, IFN-γ, and TNF-α, which play a pivotal role in controlling the growth of the parasite and its elimination (29). The protective immune responses to *Salmonella* infection are complex and involve both humoral and cellular immune responses (30,31), even though the role of humoral immunity in protection remains undefined (10). Due to the ability of *Salmonella* to persist intracellularly, cell-mediated immunity is critical to clearance of infection. In this study, the concentrations of pro and anti-inflammatory cytokines varied considerably from one group to another. IL-1β levels in the malaria mono and typhoid mono groups showed no significant variation when compared to the control but were significantly lower in these groups compared to the typho-malaria group (p<0.05) **(Figure 3b).** This suggests that co-infection triggered immune suppression in our study which is consistent with the idea that co-infection can lead to immune exhaustion, where the immune system is unable to effectively respond to one or both of the pathogens. Interleukin (IL)-1β as a pro-inflammatory cytokine has a role in disease-related inflammation, including malaria and typhoid fever (32). However, reports on the effect of IL-1β on malaria severity are inconsistent. Lyke *et al*., in 2004 (33) reported no significant change in IL-1β attributing this to the downregulation by IL-10 levels. This could also explain why the IL-1β level in this present study was highest in typho-malaria group with a lower IL-10 level. On the contrary, the level of IL-1β was significantly lower in the malaria group with a higher level of IL-10.

It has been established that during early *Salmonella* infection, inflammatory monocytes produce anti-microbial factors such as TNF-α and IL-1β (34). In the study, we observed a significant rise in TNF-α levels in the both the typhoid mono- and co-infected groups compared to the control. Unlike in the report of Lyke *et al*., in 2004 (33) who reported an insignificant difference in TNF-α level in malaria patients, we obtained a rather very high significant difference in this group compared to the control. In the malaria group, there was a strong positive correlation between IL-1β levels and TNFα (ρ=0.589, p=0.006). The concentrations of IL-2 (*p<0.001*) were also significantly upregulated in patients with malaria and typhoid comorbidity, comparing to healthy controls in which there was a weak negative correlation between IL-2 and TNF-α (ρ=-0.467, p=0.03). TNF-α elevation has been previously associated with anemia and high-density *P. falciparum* infection (35), whereas reduced IL-10 demonstrated in African children with severe malaria-induced anemia. In our study, the mean concentration of IL-10 was significantly higher in patients with malaria only, 293.12 (±106.80), compared to those with comorbidity, 255.30 (±176.95) and healthy controls 130.93 (±51.56). In the typhoid fever group, IL-2 was equally strongly and positively correlated to IL-6 (r=-0.464, p=0.01). In the malaria and typhoid fever co-morbidity group, a positive correlation was recoded between IL-2/TNFα (r=0.515, p=0.03), IL-2/IL-6 (r=0.536, p=0.02) and IL-6/TNF-α (r=0.664, p=0.007). Correlation between these cytokines suggests a complex interaction between immune cells in the malaria and typhoid fever co-morbidity which might contribute to disease severity as seen in the significant increase in TNF-α levels and the decrease in IL-10 levels in patients with malaria and typhoid comorbidity compared to those with malaria only.

In line with a previous report (36), we recorded a significant rise in IFN-γ levels in all patient groups with more significant increase in malaria patients with higher parasite densities and typho-malaria comorbidity. The level of IFN-γ in typhoid fever patient was also significantly high and confirming the report by Sheikh *et al*., in 2011 (37) confirming that IFN-γ responses against *S. Typhi* antigens are elevated in both acute and convalescent stages of human infection compared with healthy controls. Correlation analysis in this study revealed that IL-2 equally gave a strong negative correlation with IFN-γ in the typho-malaria group (r=-0.577, p<0.008). The strong negative correlation between IL-2 and IFN-γ suggests that these two cytokines have opposing effects in the immune response to typho-malaria.

The interplay between inflammatory and cortisol responses modulates an appropriate response to a stressor(11). Our study revealed a critical link between stress and the inflammatory response, particularly in the co-infected group. We observed a positive correlation between stress scores and pro-inflammatory cytokines IL-2, IFN-γ, and IL-6. This suggests that psychological stress might exacerbate the inflammatory response in patients with malaria and typhoid co-infection. A previous work revealed that, after controlling for demographic factors, a rise in IL-6 was related to increased cortisol in other conditions (11). In contrast, the correlation between perceived stress and anti-inflammatory cytokines IL-4 and IL-10 was not statistically significant in the co-infected group. A. Wolkow *et al*., in 2015 (11) observed a less pronounced association between TNF-a, IL-10, IL-4, and cortisol. This suggests that the relationship between stress and anti-inflammatory cytokines may be more complex and influenced by various factors. Further investigation is needed to elucidate the underlying mechanisms by which stress modulates the cytokine profile specifically in co-infections.

Unlike in the mono infected patients, cortisol level in the typho-malaria group showed a negative correlation with IL-6 levels (r=-0.411, p=0.03), and TNF-α levels (r=-0.413, p=0.01) suggesting a possible dampening effect of cortisol on inflammation in the co-infected group possibly due to the overwhelming immune response to the dual infection. In contrast, there was a weak positive correlation between cortisol levels and IL-4 (r=0.389, p=0.03) in the same group. The positive correlation between cortisol and IL-4 in Typho-malaria might be related to a specific immune pathway activated in response to the co-infection. Cortisol can sometimes have immunomodulatory effects, influencing the immune system in both pro- and anti-inflammatory (38) ways depending on the context. In this scenario, the initial stress response might trigger cortisol release, which then leads to some IL-4 production. However, in co-infection, this feedback loop might be dysregulated, leading to a paradoxical increase in both cortisol and IL-4. A compensatory mechanism could also occur wherein increased pro-inflammatory response due to the co-infection might trigger cortisol release to dampen inflammation. However, this might also indirectly stimulate IL-4 production as a secondary anti-inflammatory response. Data on the association between cortisol and IL-4 in typhoid fever and/or malaria is scarce. There was no significant correlation between the stress marker, cortisol and inflammatory cytokines in the typhoid mono and malaria mono-infection groups. This suggests a switch in immune response when the body deals with only typhoid or malaria. The delay in presentation for medical examination by some participants especially in the typhoid mono (mean, 4.95 days) and typhoid co-infected (mean, 4.95 days) groups may have contributed to the snapshot of cytokine patterns observed at the time of admission.

Inflammatory cytokines are essential mediators during parasitic infection and the balance between pro- and anti-inflammatory cytokines may be important for the clinical outcome of both malaria and typhoid fever. Previous reports suggest that the balance between pro- and anti-inflammatory cytokines determines parasite load and disease outcome (39–41). In contrast, other evidences suggest that disease outcome depends on cytokine overproduction and not on the balance between them, since high levels of anti-inflammatory as well as pro-inflammatory cytokines may be associated with disease severity and mortality (42). Our findings revealed a clear distinction in the immune response between the different clusters. Patients with malaria or typhoid fever alone exhibited a lower ratio of TNF-α to IL-10. This suggests a potentially well-regulated immune response, characterized by controlled inflammation to combat the infection while minimizing tissue damage. In contrast, the co-infected group displayed a significantly higher TNF-α/IL-10 ratio, indicative of a more robust and potentially detrimental inflammatory response. This heightened inflammatory state might be the body’s attempt to address the dual challenge which might lead to complications. Interestingly, the co-infection group showed a higher IL-2/IL-10 and IFN-γ/IL-10 ratios, indicating a more pronounced effort to activate T-cells and macrophages and an overreaction might potentially provoke a collateral damage. These findings collectively suggest that co-infection with malaria and typhoid fever leads to a more severe and dysregulated immune response compared to single infections. The observed imbalance between pro-inflammatory and anti-inflammatory responses in co-infection could contribute to the development of complications. Furthermore, understanding these distinct cytokine profiles may hold promise for the development of targeted immunomodulatory therapies to manage co-infections more effectively.

## Conclusion

In conclusion, our study sheds light on the intricate relationship between stress, cortisol levels, and the immune response in patients with malaria, typhoid fever, and co-infections. The findings suggest that stress may exacerbate inflammation in co-infected individuals, while cortisol may have anti-inflammatory effects in this group. The differences in cytokine profiles between mono-infected and co-infected individuals highlight the complexity of the immune response in these diseases. Moreover, exposed to severe stressors may disrupt this immune response and contribute to negative health outcomes. Our research underscores the importance of understanding the interplay between infectious diseases, stress, and the immune system for developing effective treatment strategies. Future studies should further explore the mechanisms by which stress influences cytokine expression and investigate the potential therapeutic benefits of stress management strategies in conjunction with conventional treatments for malaria and typhoid fever. Ultimately, by delving deeper into this complex relationship, we can better understand the immunopathogenesis of these diseases and improve patient outcomes.

## Data availability

The data collected in this study are available upon request to the corresponding author.

## Acknowledgments

We remain grateful to all the participants and personnel from the Obala District and Etoug-Ebe Baptist Hospitals for the collaboration. We thank the staff of the Regional Delegation of Public Health, Cameroon for the approval and monitoring of the research.

## Authors’ contributions

**Conceptualization:** Palmer Masumbe Netongo, Wilfred Fon Mbacham

**Formal analysis:** Severin Donald Kamdem, MacDonald Bin Eric, Tchoutang Ange Maxime

**Funding acquisition:** Palmer Masumbe Netongo

**Investigation:** MacDonald Bin Eric, Tchoutang Ange Maxime, Nzuno Christine

**Software:** Tchoutang Ange Maxime, Tchoupe Kamoua Eric Berenger

**Supervision:** Palmer Masumbe Netongo, Wilfred Fon Mbacham

**Writing – original draft:** Palmer Masumbe Netongo, MacDonald Bin Eric

**Writing – review & editing:** Nzuno Christine, Ngum Leslie Ngum, Bongkiyung Donald Buri, Jean Paul Chedjou, Severin Donald Kamdem, Palmer Masumbe Netongo, Wilfred Fon Mbacham, Akindeh Mbu Nji.

## References

1. Organization WH. World malaria World malaria report report. 2023.

2. Hancuh M, Walldorf J, Minta AA, Tevi-benissan C, Christian KA, Nedelec Y, et al. Typhoid Fever Surveillance, Incidence Estimates, and Progress Toward Typhoid Conjugate Vaccine Introduction — Worldwide, 2018 – 2022. 2023;72(7):171–6.

3. Ekesiobi AO, Igbodika MC, Njoku OO. CO-INFECTION OF MALARIA AND TYPHOID FEVER IN A TROPICAL COMMUNITY. Anim Res Int. 2008;5(3):888–91.

4. Netongo PM, Eric M Bin, Chedjou JP, Kamdem SD, Achonduh-atijegbe O, Mbacham WF. Malaria and Typhoid Fever Co-infection Amongst Febrile Patients in Yaoundé, Cameroon : Implication in the Genetic Diversity of Plasmodium falciparum To cite this article : 2022;7(2):47–53.

5. Wilairatana P, Mala W, Milanez GDJ, Masangkay FR, Kotepui KU, Kotepui M. Increased interleukin - 6 levels associated with malaria infection and disease severity : a systematic review and meta - analysis. Sci Rep [Internet]. 2022;1–24. Available from: 10.1038/s41598-022-09848-9

6. Id MK, Mala W, Kwankaew P, Kotepui KU, Masangkay FR, Wilairatana P. Distinct cytokine profiles in malaria coinfections : A systematic review. 2023;1–19. Available from: 10.1371/journal.pntd.0011061

7. Bradle J. TNF-mediated inflammatory disease. J Pathol. 2008;214:149–60.

8. Butler T, Ho MAY, Acharya G, Tiwari M, Gallati H. Interleukin-6, Gamma Interferon, and Tumor Necrosis Factor Receptors in Typhoid Fever Related to Outcome of Antimicrobial Therapy. 1993;37(11):2418–21.

9. Bhuiyan S, Sayeed A, Khanam F, Leung DT, Bhuiyan TR, Sheikh A, et al. Cellular and Cytokine Responses to Salmonella enterica Serotype Typhi Proteins in Patients with Typhoid Fever in Bangladesh. Am J Trop Med Hyg. 2014;90(6):1024– 30.

10. Sztein MB, Salerno-goncalves R, Mcarthur MA. Complex adaptive immunity to enteric fevers in humans : lessons learned and the path forward. Front Genet [Internet]. 2014;5(October):516. Available from: doi: 10.3389/fimmu.2014.0051

11. Wolkow A, Aisbett B, Reynolds J, Ferguson SA, Main LC. Relationships between inflammatory cytokine and cortisol responses in firefighters exposed to simulated wildfire suppression work and sleep restriction. Physiol Rep. 2015;3(11):12604.

12. Chan SF, La Greca AM. Perceived Stress Scale (PSS) [Internet]. In: Gellman, M.D., Turner, J.R. (eds) Encyclopedia of Behavioral Medicine. Springer, New York. 2013. Available from: 10.1007/978-1-4419-1005-9_773

13. Achonduh-Atijegbe OA, Mfuh KO, Mbange AHE, Chedjou JP, Taylor DW, Nerurkar VR, et al. Prevalence of malaria, typhoid, toxoplasmosis and rubella among febrile children in Cameroon. BMC Infect Dis [Internet]. 2016;16(1):1–9. Available from: 10.1186/s12879-016-1996-y

14. Nsutebu E F, Ndumbe P M AD. The distribution of anti-Salmonella antibodies in the sera of blood donors in Yaoundé, Cameroon. Trans R Soc Trop Med Hyg. 2002;96(1):68–9.

15. Noorbakhsh S, Rimaz S, Rahbarimanesh AA MS. Interpretation of the Widal Test in Infected Children. Iran J Public Health. 2003;32(1):35–7.

16. Awoke N, Arota A. Profiles of hematological parameters in Plasmodium falciparum and Plasmodium vivax malaria patients attending Tercha General. Infect Drug Resist. 2019;12:521–7.

17. Ndako JA, Dojumo VT, Akinwumi JA, Fajobi VO, Owolabi AO, Olatinsu O. Changes in some haematological parameters in typhoid fever patients attending Landmark University Medical Center, Omuaran-Nigeria. Heliyon [Internet]. 2020;6:1–5. Available from: 10.1016/j.heliyon.2020.e04002

18. Nlinwe NNO, Tang B. Assessment of Hematological Parameters in Malaria, among Adult Patients Attending the Bamenda Regional Hospital. Hindawi. 2020;1–8.

19. Kotepui M, Piwkham D, Phunphuech B, Phiwklam N. Effects of Malaria Parasite Density on Blood Cell Parameters. PLoS One. 2015;10(3):1–11.

20. Bayleyegn B, Asrie F, Yalew A, Woldu B. Role of Platelet Indices as a Potential Marker for Malaria Severity. Hindawi. 2021;1–8.

21. Lacerda MVG, Mourão MPG, Coelho HC, Santos JB. Thrombocytopenia in malaria: Who cares? Vol. 106, Memorias do Instituto Oswaldo Cruz. 2011. p. 52–63.

22. Shaikh MS, Ali B, Janjua M, Akbar A, Haider SA, Moiz B, et al. Plasmodium in the bone marrow : case series from a hospital in Pakistan, 2007 – 2015. Malar J [Internet]. 2021;20(254):1–6. Available from: 10.1186/s12936-021-03792-1

23. Baffour SA, Mensah BT, Johnson G, Naa D, Armah O, Mustapha SA, et al. Haematological parameters and their correlation with the degree of malaria parasitaemia among outpatients attending a polyclinic. Malar J [Internet]. 2023;22:1–8. Available from: 10.1186/s12936-023-04710-3

24. Ibrahim EA, Kheir MM, Elhardello OA, Almahi WA, Ali NI, Elbashir MI IA. Cortisol and uncomplicated Plasmodium falciparum malaria in an area of unstable malaria transmission in eastern Sudan. Asian Pac J Trop Med. 2011;146–7.

25. Vedantam D, Poman DS, Motwani L, Asif N, Patel A, Anne KK. Stress-Induced Hyperglycemia: Consequences and Management. Cureus. 2022;1(7):14.

26. Whitworth JA, Williamson PM, Mangos G, Kelly JJ. Cardiovascular consequences of cortisol excess. Vasc Health Risk Manag. 2005;1(4):291–9.

27. Glaser R, Kiecolt-Glaser JK. Stress-induced immune dysfunction: implications for health. Nat Rev Immunol [Internet]. 2005;5(3):243–51. Available from: 10.1038/nri1571

28. Sandeep BR, Bhutto MG, P SBK. Study of serum cortisol levels in complicated and uncomplicated Plasmodium vivax malaria patients. Int J Res Med Sci |. 2021;9(1):254–61.

29. Popa GL, Popa MI. Recent Advances in Understanding the Inflammatory Response in Malaria : A Review of the Dual Role of Cytokines. J Immunol Res. 2021;2021:10–2.

30. Sztein MB. Cell-mediated immunity and antibody responses elicited by attenuated Salmonella enterica serovar Typhi strains used as live oral vaccines in humans. Clin Infect Dis. 2007;15–9.

31. Levine MM, Tacket CO, Sztein MB. Host – Salmonella interaction : human trials. Microbes Infect. 2001;3:1271–9.

32. Mahittikorn A, Kwankaew P, Rattaprasert P, Kotepui KU. Elevation of serum interleukin - 1β levels as a potential indicator for malarial infection and severe malaria : a meta - analysis. Malar J [Internet]. 2022;1–11. Available from: 10.1186/s12936-022-04325-0

33. Lyke KE, Burges R, Cissoko Y, Sangare L, Dao M, Diarra I, et al. Serum Levels of the Proinflammatory Cytokines Interleukin-1 Beta ( IL-1 ␤ ), IL-6, IL-8, IL-10, Tumor Necrosis Factor Alpha, and IL-12 ( p70 ) in Malian Children with Severe Plasmodium falciparum Malaria and Matched Uncomplicated Malaria or Healthy Co. Infect Immun. 2004;72(10):5630–7.

34. Pham OH, McSorley SJ. Protective host immune responses to Salmonella infection. Natl Inst Heal. 2015;10:101–10.

35. Shaffer N, Grau GE, Hedberg K, Davachi F, Lyamba B, Hightower AW, et al. Tumor necrosis factor and severe malaria. he J Infect Dis. 1991;1(163):96–101.

36. Oyegue-liabagui SL, Bouopda-tuedom AG, Kouna LC, Maghendji-Nzondo S, Nzoughe H, Tchitoula-makaya N, et al. Pro- and anti-inflammatory cytokines in children with malaria in Franceville, Gabon. Am J Clin Exp Immunol. 2017;6(2):9– 20.

37. Sheikh A, Khanam F, Sayeed A, Rahman T, Pacek M, Hu Y, et al. Interferon-gamma and Proliferation Responses to Salmonella enterica Serotype Typhi Proteins in Patients with S. Typhi Bacteremia in Dhaka, Bangladesh. PLoS Negl Trop Dis. 2011;5(6):1193.

38. Coutinho AE, Chapman KE. The anti-inflammatory and immunosuppressive effects of glucocorticoids, recent developments and mechanistic insights. Mol Cell Endocrinol. 2011;335(1):2–13.

39. Winkler S, Willheim M, Baier K, Schmid D, Aichelburg A, Graninger W, et al. Reciprocal Regulation of Th1- and Th2-Cytokine-Producing T Cells during Clearance of Parasitemia in Plasmodium falciparum Malaria. Infect Immun. 1998;66(12):6040–4.

40. Sarangi A, Mohapatra PC, Dalai RK, Sarangi AK. Serum IL-4, IL-12 and TNF-alpha in malaria : a comparative study associating cytokine responses with severity of disease from the Coastal Districts of Odisha. J Parasit Dis. 2014;38(2):143–7.

41. Punnath K, Dayanand KK, Chandrashekhar VN, Achur RN, Kakkilaya SB, Ghosh SK, et al. Association between inflammatory cytokine levels and anemia during Plasmodium falciparum and Plasmodium vivax infections in Mangaluru: A Southwestern Coastal Region of India. Trop Parasitol. 2019;9(2):98–107.

42. Day NP, Hien TT, Schollaardt T, Loc PP, Chuong L V., Chau TT, et al. The prognostic and pathophysiologic role of pro-and antiinflammatory cytokines in severe malaria. J Infect Dis. 1999;180(4):1288–1297.

